# Pediatric surgical services in Bangladesh during the COVID 19 pandemic: How they are affected and how to overcome the backlog, keeping healthcare professionals safe

**DOI:** 10.1101/2020.08.16.20169979

**Authors:** Md Jafrul Hannan, Mosammat Kohinoor Parveen, Md Samiul Hasan

## Abstract

**Background:** Severe Acute Respiratory syndrome coronavirus 2 (SARS-CoV-2), which originated in Wuhan, China, has turned into a pandemic. All countries have implemented multiple strategies to try mitigating the losses caused by this virus. To stop the rapid spread of the disease and in compliance with the World Health Organization’s social distancing policy, the government of Bangladesh has implemented a number of strategies, one of which is to limit the spread of the virus in hospitals by postponing elective procedures and providing only emergency services in the hospitals.

The objective of this survey was to assess the current status of pediatric surgical procedures in different hospitals in Bangladesh and assess the effects of the current restrictions along with their implications in the long run.

**Materials and Methods:** A survey was performed among doctors from public and private hospitals in Bangladesh to evaluate the status on pediatric surgery.

**Results:** The results clearly revealed the lack of a significant reduction in doctors’ exposure to SARS-CoV-2 by postponing elective procedures.

**Conclusion:** Keeping in mind the socioeconomic and health care conditions of the country, the author recommend resuming elective surgical procedures.

## Introduction

The Coronavirus Disease 2019 (COVID-19) has shaken the world to its core. The causative agent, severe acute respiratory syndrome coronavirus (SARS-CoV-2), emerged in China in December 2019 and spread rapidly worldwide.[1] On March 11, 2020, the World Health Organization (WHO) declared COVID-19 a pandemic.[2] Since then, it has led to a significant disturbance in the health sector, economy, and quality of life. Most countries have taken and implemented numerous initiatives to control the spread of the virus. Despite these efforts, people keep becoming infected. As of July 22, 14,765,256 people had been infected with SARS-CoV-2 globally, and among these, 612,054 died.[3]

In compliance with the social distancing measures recommended by the WHO, healthcare institutions worldwide have taken several initiatives to limit non-essential hospital visits. [4] These include encouraging telehealth for less severe health problems, prioritization of severe health problems, providing treatment for emergency life-threatening conditions only, and delaying elective procedures as much as possible. The goal is to control the pandemic and ensure the necessary treatment at the same time.

These control measures have resulted in an unexpected strain on the health care system, especially on surgical services. It is still not clear when restrictions will be lifted and how to reschedule the postponed cases once the pandemic is gone. Numerous protocols and strategies have been described to maintain the flow of pediatric surgical care, prevent the spread, and to protect the health care workers at the same time.[5-8] As of July 20, more than 2 million confirmed cases have been reported in Bangladesh, with a case fatality rate of 1.29%.[9] Unfortunately, 11% of the confirmed cases are healthcare workers; therefore, there is a growing fear among the healthcare workers actively participating in the fight against COVID-19.[10] Given these circumstances, we intended to evaluate the condition of pediatric surgical services in Bangladesh during the COVID 19 pandemic.

## Materials and Methods

For the purpose of evaluating the pediatric surgical services in the country, a survey was carried out that included doctors working in both public and private hospitals. The survey was carried out to know about the period starting from April 1, 2020 until June 30, 2020. The survey involved doctors of varying age and of both sexes working in Pediatric Surgical Departments, addressed the infection status of the patients who were operated upon; i.e., whether they were SARS-CoV-2-positive or not. The respondents were queried if they or any of their team members at work became SARS-CoV-2-positive during the period, were also asked if they had been noticing any reduction in workload related to surgical services as compared with pre-COVID-19 levels. The last question of the survey was focused on whether the respondent felt mentally prepared to commence performing routine operations at a level commensurate to that of before COVID-19 given the current situation.

## Results

The survey was responded by 52 doctors including young professionals from the age of 25 and one respondent above the age of 65. Out of these 52 respondents, only one had not performed any emergency surgery whereas, 8 out of the 52 respondents responded that they did not perform any routine surgery during the same period (Table 1). Emergency operations were done by all teams during this period (Supplemental data).

**Table 1:** How many routine operations performed by the team

| ANSWER CHOICES | RESPONSES |
| --- | --- |
| 00 | 15.38% (8) |
| 1-9 | 38.46% (20) |
| 10-19 | 26.92% (14) |
| 20-29 | 7.69% (4) |
| 30-39 | 3.85% (2) |
| 40-49 | 1.92% (1) |
| 50 and above | 5.77% (3) |
| TOTAL | 52 |

Around 65% of the participants responded that at least one of their patients had been suspected of having COVID-19 (Fig. 1). About 50% of the respondents replied that none of their team members became Covid-19 positive during study period that means at least 50% contracted the virus. About 75% of the respondents answered that there had been a decline of up to 70% in the number of surgical procedures as compared with pre-COVID-19 levels, and almost all the respondents had noticed a decline in the number of surgeries they had performed (Figure 2). Only a third of the respondents has agreed to resume routine operations like that before Covid-19 (Supplemental data).

**Figure 1:**
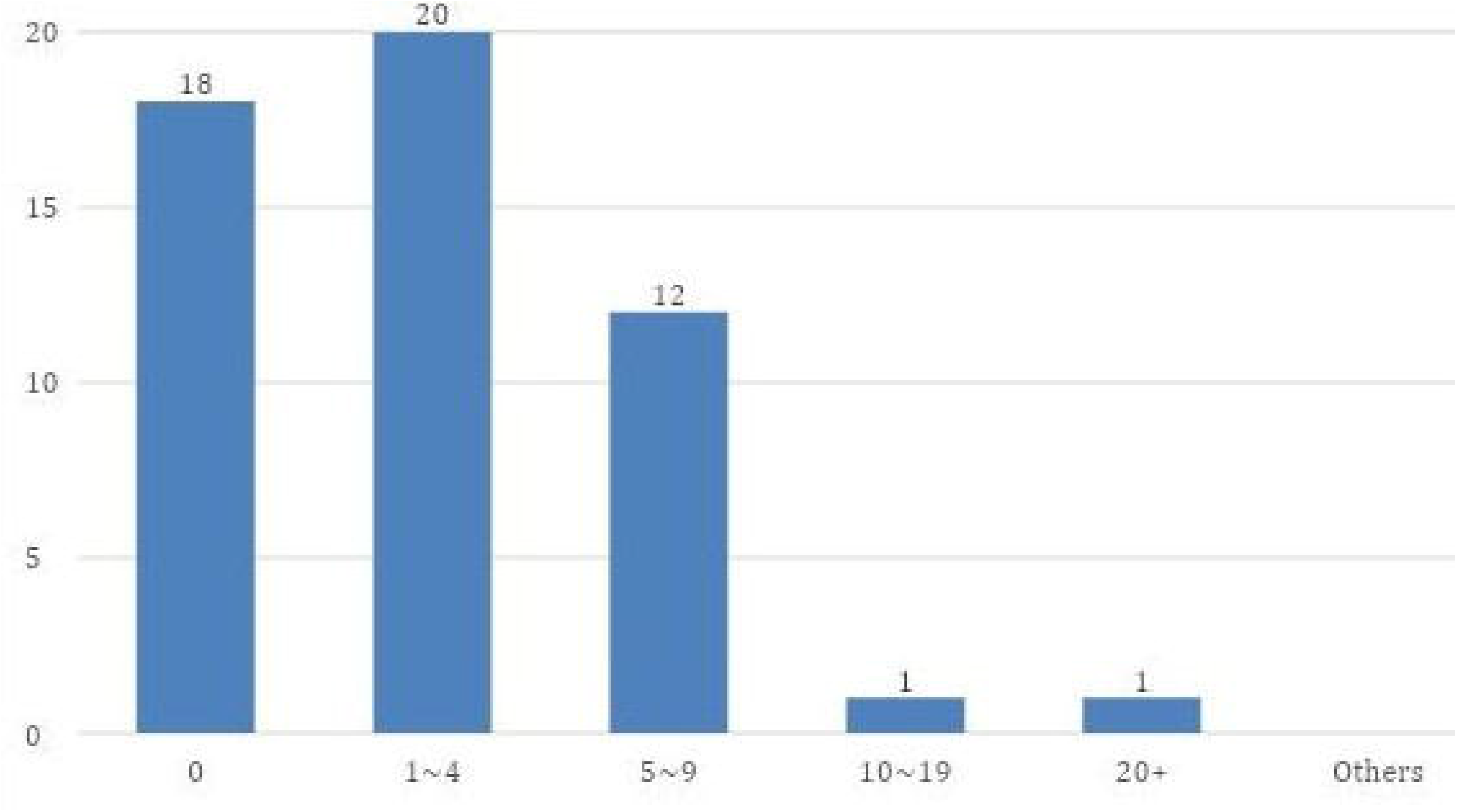
Number of Covid-19 patients operated by a team.

**Figure 2:**
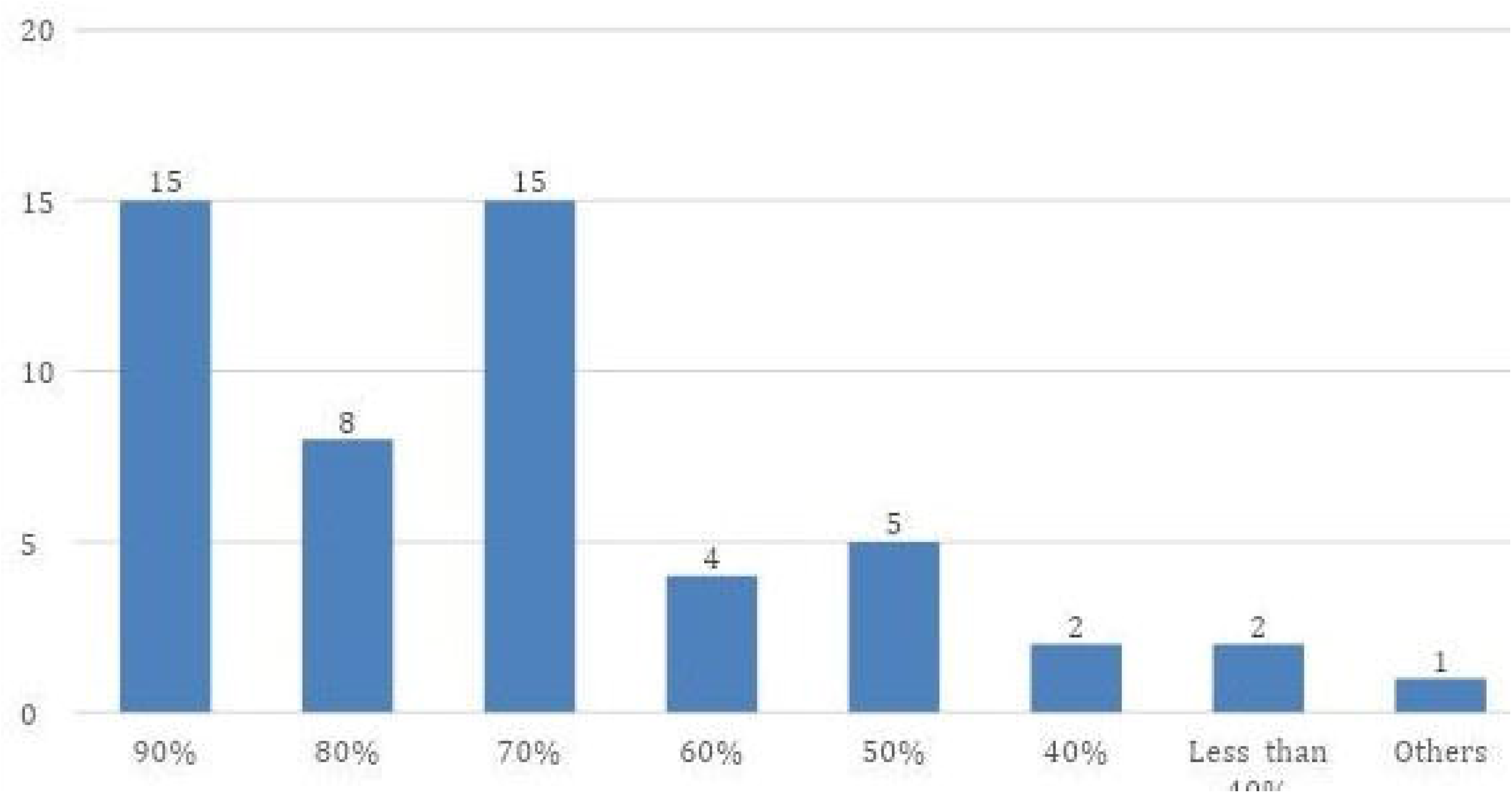
Percentage of volume of operation reduced in comparison to pre-Covid-19 period.

From the survey it is evident that there has been a drastic reduction in the number of surgeries performed now and during pre COVID-19 times. Also, around 50% of the respondents replied that none of their team members or them contracted COVID-19 during the period but 50% responded having contacted the virus. Also, 1 out 52 respondents and 8 out of 52 respondents were not performing emergency and routine surgeries respectively (Supplemental data).

## Discussion

It is evident from the results of the survey that delaying elective surgical procedures is not having any significant effect in terms of reducing the number of doctors that become SARS-CoV-2-positive. In the setting of a developing country the question remains how far it is justifiable to not utilize an infrastructure and workforce that is already far behind the standard requirements. [11] The strategy of withholding routine operations in the initial stages of the pandemic was mostly based on lack of knowledge rather than fact. There is evidence that the biggest manufacturing industry in Bangladesh (Garments) remained open since mid-April, which did not result in an SARS-CoV-2 infection rate among garment workers beyond the rate observed in the general population. Shutting down healthcare facilities, particularly in case of pediatric surgery, might not have been a useful decision, since children are less affected by COVID-19.[12] At the same time, this decision might have triggered anxiety among the general population, which has led them to abstain from going to hospitals even for emergency surgical conditions, resulting in disease complications. The withholding of routine operations has already produced a large backlog in the setting of an overloaded workforce, which may take more than a year to clear up.

## Conclusion

Based on the data from the present survey, the recommendation would be to resume full-fledged pediatric surgical services in Bangladesh as well as in countries with similar socioeconomic status. In future similar situations it would be advisable to not stop usual routine surgical services completely.

## Data Availability

All relevant data were included in the manuscript. Additional data are available in supplemental data.

## Contributors

**Md Jafrul Hannan:** Conception, study design, data collection, data analysis, manuscript writing & editing.

**Mosammat Kohinoor Parveen:** Data analysis, and manuscript revision.

**Md Samiul Hasan:** Manuscript writing. Editing, and submission.

